# Associations between 40-year trajectories of BMI and proteomic and epigenetic aging clocks: deciphering nonlinearity and interactions

**DOI:** 10.1101/2025.03.21.25324375

**Authors:** Gabin Drouard, M. Austin Argentieri, Aino Heikkinen, Miina Ollikainen, Jaakko Kaprio

**Affiliations:** Institute for Molecular Medicine Finland (FIMM), HiLIFE, University of Helsinki, Helsinki, Finland; Analytic and Translational Genetics Unit, Massachusetts General Hospital, Boston, MA, USA; Program in Medical and Population Genetics, Broad Institute of MIT and Harvard, Boston, MA, USA; Minerva Foundation Institute for Medical Research, Helsinki, Finland

**Keywords:** Biological aging, Proteomics, Epigenetics, Weight change, Body mass index, Obesity, Nonlinearity, Interactions

## Abstract

**Background:** While studies have examined associations between changes in BMI and biological aging, the use of biological age estimates derived from omics other than DNA methylation data as well as nonlinearity and interactions in these associations are underexplored.

**Objective:** We aimed to investigate how BMI at ages 18 and ∼60, as well as changes in BMI from age 18 to ∼60, relate to downstream epigenetic and proteomic aging. We also examined nonlinearity and interactions in these associations.

**Methods:** We analyzed data from 401 Finnish participants with up to 9 self-reported or measured BMI values collected over 40 years. Olink proteomics and Illumina DNA methylation data were generated from blood samples taken at the last BMI measurement. We calculated 4 and 5 estimates of biological age from proteomic and epigenetic clocks, respectively. Changes in BMI over time were estimated using mixed-effects models. We applied generalized additive models to explore 1) nonlinearity in associations between BMI trajectories and biological aging while adjusting for chronological age and 2) smooth interactions between baseline BMI with changes in BMI and BMI at ∼60 years old.

**Results:** BMI at 18 and ∼60 years old and changes in BMI were associated with increased biological aging for most aging estimates. We found statistical evidence of nonlinearity for about one-third of the significant associations, mostly observed for proteomic clocks. We identified suggestive evidence for interactions between BMI at 18 years and BMI at ∼60 years in explaining variability in two proteomic clocks (p=0.07; p=0.09).

**Conclusion:** Our study illustrates the potential of proteomic clocks in obesity research and highlights that assuming linearity in associations between BMI trajectories and biological aging is a critical oversight. Associations between BMI and biological aging are likely modulated by past BMI, which warrants validation by other studies.

## Introduction

Obesity is a major public health problem and its incidence is increasing worldwide [1]. Importantly, individuals with obesity tend to remain obese, regardless of multiple attempts to lose weight, highlighting the urgent need for early prevention efforts [2]. While weight loss is a major focus of the scientific community, understanding the correlates and etiology of long-term weight gain may allow for targeted prevention before obesity develops. Meanwhile, aging is a major risk factor in predicting many diseases [3], and both obesity and aging interact in predicting mortality [4]. Studies have also suggested that obesity may cause changes in aging of the cellular systems [5,6], consequently leading to an increased risk of age-related diseases, reinforcing the need to disentangle the underpinnings of the association between obesity and aging.

In recent years, biological aging has emerged as a new avenue for aging research [7,8] and could serve as a new variable of interest in assessing how body weight is biologically related to aging. Biological aging estimates aim to assess an individual’s body age, whereas chronological age represents an individual’s age calculated as the time elapsed since birth. Biological aging, therefore, by definition, is a better reflection of an individual’s physiological state than chronological age and better captures the influences of lifestyle factors, such as diet or physical activity, on aging. When biological age is greater than chronological age, individuals are said to have accelerated biological age, which is known to be a good predictor of onset and development of multiple diseases [9]. Several so-called aging clocks have been developed to estimate biological age using DNA methylation (i.e., epigenetic) data, with Horvath [10] and GrimAge/GrimAge2 [11] clocks being among the most widely used. As DNA methylation is strongly imprinted by genetics as well as by short- and long-term environmental exposures, and is known to be partially reversible, it is a valuable resource for assessing both current and past environmental exposures on the body. It can thus be used to estimate biological age [12,13]. Recently, the development of biological age estimates using other omics such as proteomics has emerged in the literature [14,15], but their advantages over DNA methylation-based biological age estimates remain largely unknown and underexplored, especially since assessing the superiority of biological age estimates is likely to be highly context dependent.

The literature is replete with studies demonstrating associations between markers and determinants of body composition, including diet, weight, substance use and BMI, and biological aging across multiple age groups [16–23]. These studies have shown greater biological age in individuals with poor diet and lifestyle or higher BMI, as well as the effect of genotypes on the associations between biological aging BMI, metabolic health, and diet [16–18]. Longitudinal intervention studies examining the effects of dietary changes on biological aging have also shown decelerated biological aging in individuals who adopted healthier diets [20,24,25]. On the other hand, cohort studies have allowed the examination of associations between changes in weight or BMI and biological aging over relatively short to longer follow-up periods [22,26,27]; all have shown positive associations between weight gain and accelerated biological aging. As a result, there is already evidence in the literature for the longitudinal associations between weight or dietary changes and biological aging.

Although studies investigating associations between changes in body weight and biological aging have proliferated in recent years, significant challenges remain, four of which we identify in this paragraph. First, most longitudinal studies examining the associations between weight gain and biological aging are limited by relatively modest follow-up periods, whereas studies tracking anthropometric measures over several decades and across age groups could provide valuable insights for early prevention of obesity-related health risks. A second challenge is to understand how different aging clocks calculated from omics other than DNA methylation data might be related to changes in body weight, especially since the literature on connections between obesity or nutrition and biological aging is largely based on epigenetic aging clocks. To our knowledge, a comparison of epigenetic aging clocks with other biological aging clocks, such as proteomic aging clocks, in studying body weight in cross-sectional and longitudinal settings is lacking in the literature. Such results would allow the scientific community to gain a better biological and etiological understanding of how BMI and weight change translates into accelerated aging. A third challenge is to explore nonlinear patterns in associations between body weight changes and biological aging.

Exploring nonlinearity in these associations is likely to provide a more complete, holistic picture of whether variability in biological aging is better captured by recent or past body weight, as it is likely that changes in body weight influence downstream faster aging unevenly over time. As such, linear modeling would assume that one unit change in weight is associated with the same magnitude of change in biological age across adult age, which is unlikely to occur in practice. Thus, there is a need to use nonlinear models, which appear to be largely underutilized in the literature. To date, only a few studies have reported “U-shaped” patterns in associations between biological age estimates, as assessed with epigenetic aging clocks, and BMI [20,21]. Finally, whether associations between changes in body weight and biological aging vary across baseline BMI values is underexplored. Models incorporating interaction terms could provide answers to whether effects of changes in BMI on biological aging are increased in individuals with a high baseline BMI, independent of the individual effect of baseline BMI on biological aging.

Our study aims to address the aforementioned challenges through establishing associations between long-term weight change with biological aging, estimated from epigenetic or proteomic data. To do so, we analyzed a deeply phenotyped sample of same-sex twin pairs born between 1945 and 1957, first assessed in 1975. For these twins, up to nine BMI measurements were collected from their 20s to 60s, and epigenetic and proteomic age estimates were generated at the last follow-up.

Nonlinearity in associations was assessed, as well as interactions between 1) baseline BMI with changes in BMI and 2) baseline BMI with the last BMI measurement in capturing variability in biological aging. We provide a complete description of how each biological aging clock associates with BMI and its changes over time, present evidence for nonlinearity in these associations, and discuss how weight change relates to biological aging as estimated by various clocks, whether epigenetic-or proteomic-based.

## Methods

### Cohort

The current study uses a sample of twins from the older Finnish Twin Cohort (FTC) [28], called the Essential Hypertension Epigenetics (EH-Epi) sample. The EH-Epi twins were selected based on a questionnaire in 2011, and twin pairs showing discordance in their self-reported blood pressure were invited to a one-day in-person visit in Helsinki. The twins’ blood pressure was measured, blood samples were taken, and interviews were conducted as described elsewhere [29]. Multiple omics were generated from these blood samples, from which epigenetic and proteomic data were used in the current study. At the time of blood sampling, the twins were on average 62 years old (range: 56-70). Anthropometric measurements were collected at the time of blood sampling by trained research nurses, who collected information on their health status including blood pressure measures, lifestyle and current medications. In addition, self-reported weight and height were available from postal questionnaires collected in 1975, 1981, 1990 and 2011. Details about weight, height, and resulting BMI measurements are provided below.

### Anthropometric measurements

At the time the twins provided the blood samples from which the omics data were derived, their weight, height and waist circumference were also measured. Being the fifth point of measurement, we refer to the time of blood sampling as wave 5. The twins’ BMI was calculated as the ratio of their measured weight (in kg) to their squared height (in meters) and was available for all 401 twins included in the current study at wave 5. The waist to height ratio was also computed.

Additional weight and height measurements were collected from postal questionnaires sent in 1975, 1981, 1990 and 2011, i.e. waves 1 to 4. The reliability of BMI values from self-reported measures of weight and height from the wave 4 (in 2011) questionnaire were compared to the measured values (at wave 5) taken on average two years later. As reported earlier, the correlations between self-reported and measured BMI were high (r=0.95) for both men and women [30]. In 1990, in addition to their current weight, the twins reported their weight when they were 20 and 30 years old, as well as their weight 5 years before completing the questionnaire. In 2011, in addition to their current weight, the twins reported their weight 5 years before completing the questionnaire and their weight at age 50. All corresponding BMI values were calculated, with about 70% of the twins included in the current study having complete BMI information (i.e., nine complete BMI values over around 40 years of follow-up) (Figure S1). Individual missing BMI data points were not imputed but kept as missing in the statistical analyses.

### Proteomic data processing & proteomic aging clocks

Proteomic data were generated from blood plasma samples of EH-Epi twins and analyzed by an antibody-based technology based on proximity extension assay (Olink Proteomics AB, Uppsala, Sweden). Olink Explore 3072 was used to quantify assays belonging to cardiometabolic, inflammation, neurology and oncology panels. A full description of the data preprocessing has been described elsewhere [31], resulting in complete, processed, quality-controlled proteomic data for 401 twins (including 196 complete twin pairs). Briefly, sample outliers were excluded, Olink’s internal quality control criteria were verified, proteins with more than 20% of values below the limit of detection (LoD) were excluded, and values below the LoD for the remaining proteins (representing less than 1% of total data points) were replaced with the plate-specific LoD. A total of 2,321 plasma proteins were available for these twins, from which biological age was estimated as described later.

We calculated estimates of two proteomic aging clocks known as Proteomic Aging Clock (PAC) [15] and Healthspan Proteomic Score (HPS) [32] from the corresponding summary statistics. Although HPS is not intended to characterize biological aging but rather healthspan, its measure is reflective of aging; we therefore included HPS in the analyses and referred to it as an aging clock for ease of reading. Because we initially discarded proteins with high missing rates (>20%) during data processing, four and six proteins were unavailable in the calculation of PAC and HPS, respectively. We therefore calculated PAC and HPS ensuring that the sum of absolute weights across all coefficients remained unchanged, and both estimates showed moderate Pearson correlations with chronological age (PAC: r=0.48; HPS: r=-0.38). As a sensitivity analysis, we examined whether keeping the few proteins with high missing rates (>20%) in the calculation of PAC and HPS would lead to substantial differences in their correlations with chronological age. The correlations remained of similar magnitude (PAC: r=0.48; HPS: r=-0.37), suggesting that the missingness does not have an impact on the age or healthspan estimates. For ProtAge [14], we first re-normalized the expression of each protein in the EH-Epi twins data using an adapted version of the normalization procedure used in the original ProtAge paper, where protein expression was aligned with the population reference values from the UK Biobank (UKB) data used to train the model (since the LightGBM model used is sensitive to the scale of the input features). Each protein was min-max scaled to be between 0 and 1 using the trained scikit-learn *MinMaxScaler* () object from the UKB data and then centered on the median of that protein in the UKB. Seven proteins were present in the original ProtAge model but not available in the EH-Epi twins data (CXCL14, GIP, FSHB, TSPAN1, PSPN, KLK3, INSL3), which were set to NA and included in the model. Re-normalized expression data were then passed through the published ProtAge LightGBM model to produce protein predicted age estimates.

Finally, we calculated Adipose age, a recently published organ-specific proteomic clock that aims to estimate biological age from blood proteins to reflect the age of adipose tissue [33]. In the current investigation, we refer to this clock as the Adipose clock. A description of the proteomic clocks is available in the supplementary material (Table S1). Pearson correlation coefficients between PAC, HPS, ProtAge and Adipose together and with epigenetic age estimates (un)adjusted for chronological age are available in the supplementary material (Additional file 1, Table S2, Table S3; Figure S2).

### Epigenetic data processing & epigenetic aging clocks

Of the 401 twins included in the current study, 379 also had blood DNA methylation data available, allowing us to calculate their epigenetic ages. DNA methylation levels were quantified using the Infinium Illumina HumanMethylation450K array and pre-processed and normalized using the R package *meffil* [34], as described in detail elsewhere [35,36]. We calculated epigenetic ages using five different algorithms. Horvath [10], Hannum [37], and PhenoAge [38], were calculated using their PC score version [39], as described elsewhere [40]. We also calculated biological ages using GrimAge2 [11] and the pace of aging using the DunedinPACE [41] clocks. Although DunedinPACE represents the pace of aging rather than biological aging, we refer to it as an aging clock for ease of reading. A description of the epigenetic clocks, as well as their pairwise correlations together or with proteomic clocks, unadjusted or adjusted for chronological age, is available in the supplementary material (Table S1; Table S2; Table S3; Figure S2).

### Statistical analyses

#### Linear mixed effects modeling for BMI trajectory analysis

We summarized the twins’ BMI trajectories into three measures, one of which was the most recent BMI, measured at the time of blood sampling, i.e., BMI at wave 5. The other two measures were BMI at baseline and the rate of change in BMI over time. We used linear mixed effects (LME) models to obtain these two measures by deriving intercept (i.e., fitted baseline) and slope (i.e., fitted rate of change in BMI) values for everyone. LME models are flexible models for longitudinal analyses incorporating both fixed and random effects, the latter of which we used for both the intercept and slope to derive estimates that are unique to each individual. We set the age for the intercept at 18 years, so that the baseline BMI in subsequent analyses is that expected at 18 years. This choice was validated by the fact that 90.3% of the twins in our sample had available BMI measurements from age 18 to age 25 (in 1975), ensuring that the fitted baseline BMI reflected the observed BMI trajectories. LME models were fitted to allow for missing BMI values, so that individual intercept and slope values were fitted based only on the available measurements for each individual. These analyses were performed using the R package *lme4* version 1.1-30.

In addition, we tested whether linear modeling of BMI trajectories adequately described observed trajectories of BMI by 1) checking the conditional R^2^ obtained after fitting LME and 2) comparing our model with a model that included a quadratic term to account for potential nonlinearity in BMI trajectories. The conditional R^2^ was 90%, indicating relatively good model-data fit. Although the Akaike information criterion (AIC) of the linear model was greater than that of a model that included a quadratic term, which would indicate better model-data fit when using a linear-quadratic model, the quadratic term was not found to be significant (coefficient: 0.018; p-value: 0.37), indicating that at the population level, the trajectories of BMI do not show a clear quadratic trend, which is also illustrated in Figure S1. Therefore, the linear modeling of BMI trajectories was retained.

### Generalized additive models: associations and interactions

We used generalized additive models (GAMs) to examine the associations between three measures of BMI trajectories — baseline BMI, BMI at blood sampling, and changes in BMI — and estimates of biological aging. GAMs are a flexible class of statistical models that extend linear models to handle nonlinear relationships and allow the shape of the relationship between predictors and outcomes to be strongly determined by the data [42]. This is achieved by modeling outcomes as the sum of smooth functions of predictors, often implemented using splines, such as cubic or thin-plate regression splines, the latter we used in the current study. The effective degrees of freedom (edf) is a commonly used measure to assess the degree of non-linearity in association in GAMs. As such, edf quantifies the complexity or flexibility of the smooth functions used to model the relationships between predictors and outcome. The higher the edf, the greater the complexity; edf values close to one indicate a linear association (assuming the association is significant).

We fitted several GAMs with estimates of biological age as outcomes. Biological age estimates were first adjusted for chronological age and resulting residuals were scaled to facilitate downstream comparisons between biological clocks. We used rank-based inverse normal transformation as a scaling method instead of classical standardization to ensure that all outcomes were perfectly normally distributed. This then systematically satisfies one assumption of GAMs that require a Gaussian distribution of outcomes and enables downstream comparisons between aging clocks.

First, we independently assessed whether baseline BMI, BMI at blood sampling, or changes in BMI were associated with biological aging outcomes; p-values testing the null of F-statistic values were considered significant if below 0.05. For associations with changes in BMI, we also added fitted baseline BMI as a covariate to account for the fact that changes in participants’ BMI may only be informative if one recognizes that not all participants had the same BMI at baseline. We added family identifiers as random effects to ensure that family relatedness was corrected for. We then fitted counterpart models with only linear terms and compared GAMs that allowed for nonlinearity with these linear counterpart models using the AIC. We used ΔAIC, defined as the AIC of the nonlinear model minus the AIC of the linear counterpart, to indicate whether associations were nonlinear; a negative ΔAIC value indicates that a GAM assuming nonlinearity statistically improves model performance over a model assuming linearity. Analyses were conducted in R using the *mgcv*package version 1.8-39.

Finally, we sought to assess whether baseline BMI interacted with changes in BMI or BMI at blood sampling in capturing variability in biological aging (adjusted for chronological age and scaled). We modeled smooth interactions in both scenarios using two methods available as part of the *mgcv*R package: *te()*and *ti()*. Smooth interactions capture relationships between variables, allowing their combined effect to vary flexibly across their values, without being constrained by linear assumptions. While *te()*allows to evaluate the joint effect of two predictors, including their individual effects and interactions, *ti()* allows to evaluate the interaction term independently of the individual effects of the predictors. We used *te()*to assess the extent to which smooth interactions between baseline BMI and changes in BMI or BMI at wave 5 are jointly associated with biological aging, as well as the edf of this joint interaction. Evidence for nonlinearity was determined using ΔAIC as described previously. In addition, we separated predictors to their interaction using *ti()* to test for interactions between predictors independent of their respective individual effects on biological aging variability.

## Results

### Participant characteristics and BMI trajectories

A description of the participants at wave 5 is given in Table 1. The twins were 56-70 years old at wave 5 (mean: 62; SD: 3.8), and 22% of them had a BMI above 30 kg.m^-2^. On average, the twins gained 0.14 BMI units per year (SD: 0.08; interquartile range: 0.08-0.19) over the 40-year follow-up, which corresponds to a weight gain of 450 g per year for a person of 1.80 m in height. Females and males had the same average change in BMI over time (0.14 units/year), but with slightly greater variability in females (SD: 0.09) than in males (SD: 0.07). Correlations between biological age estimates and chronological age ranged 0.11-0.77 in absolute values in EH-Epi, with Adipose and ProtAge showing the lowest and strongest correlations, respectively (Table S2, Figure S2). Pairwise correlations between biological aging clocks are also available in the supplementary material (Table S2, Table S3).

**Table 1:** Description of EH-Epi twins at last measurement (i.e., at blood sampling; wave 5). IQR: Interquartile range. N: Number of twins for which information is available. N_cases_: Number of twins with a particular binary trait. SD: Standard deviation.

|  |  |  | mean or N<sub>cases</sub> | SD or % | IQR | Range |
| --- | --- | --- | --- | --- | --- | --- |
| Continuous variables | Alcohol consumption (g/months) | 387 | 321 | 429.9 | 70.2 - 385.5 | 0.0 - 4928.0 |
|  | Body mass index (kg.m-2) | 401 | 27.3 | 4.9 | 24.0 - 29.6 | 18.1 - 46.1 |
|  | Waist circumference (cm) | 401 | 94.4 | 14.6 | 84.5 - 103.0 | 56.0 - 140.0 |
|  | Waist-to-height ratio | 401 | 0.56 | 0.08 | 0.51-0.61 | 0.37 - 0.89 |
|  | Systolic blood pressure (mmHg) | 401 | 143.3 | 16.9 | 131.5 - 154.5 | 104.2 - 217.5 |
|  | Diastolic blood pressure (mmHg) | 401 | 83.5 | 10.1 | 77.2 - 89.5 | 58.5 - 123.2 |
| Binary variables | Females | 401 | 237 | 59% |  |  |
|  | Never smokers | 398 | 188 | 47% |  |  |
|  | Former smokers | 398 | 146 | 36% |  |  |
|  | Daily smokers | 398 | 53 | 13% |  |  |
|  | Antihypertensive medication | 401 | 173 | 43% |  |  |

### Associations between BMI trajectories and biological age acceleration

We investigated whether biological aging, as assessed by 9 different aging clocks adjusted for chronological age, was associated with BMI trajectories of the twins over four decades of follow-up. GAMs modeled scaled biological age (adjusted for chronological age) as the outcome. Baseline BMI was significantly positively associated with all biological age estimates, adjusted for chronological age, except for the Adipose clock (Figure S3; Table 2). The explained deviation was at least 10% for two of these aging clocks: HPS and GrimAge2. We found evidence that two clocks, PAC and GrimAge2, were nonlinearly associated with baseline BMI. No evidence for nonlinearity in associations with the other estimates of biological aging was observed.

**Table 2:** Associations between trajectories of BMI and aging clocks as assessed with generalized additive models. Aging clock estimates were adjusted for chronological age and residuals were scaled. In models assessing the association between changes in BMI and biological aging, baseline BMI was added as a covariate, but model performance metrics (R^2^, %Dev) exclude the effect of baseline BMI in the model. ΔAIC is defined as the AIC of the nonlinear model minus the AIC of the linear counterpart, to indicate whether associations were nonlinear. %Dev: percentage of explained variation. R2: coefficient of determination. edf: effective degrees of freedom.

| Predictor | Aging clock | edf | F-statistic |  | GAM performance |  |  |
| --- | --- | --- | --- | --- | --- | --- | --- |
| | | | F value | p-value | %Dev | R2 | Best fit ($\Delta AIC$) |
| Baseline BMI | PAC | 2.45 | 12.4 | <1.0E-16 | 9.3 | 8.7% | linear (>0) |
| Baseline BMI | HPS | 3.16 | 14.5 | <1.0E-16 | 13.3 | 12.6% | nonlinear (<0) |
| Baseline BMI | ProtAge | 1.87 | 5.9 | 2.0E-03 | 3.6 | 3.2% | linear (>0) |
| Baseline BMI | Adipose | 1.32 | 1.0 | 2.5E-01 | 0.7 | 0.4% | None |
| Baseline BMI | Horvath | 1.06 | 14.1 | 1.1E-04 | 4.1 | 3.9% | linear (>0) |
| Baseline BMI | Hannum | 1.07 | 15.8 | 4.0E-05 | 4.7 | 4.5% | linear (>0) |
| Baseline BMI | PhenoAge | 1.00 | 23.1 | 2.3E-06 | 5.8 | 5.5% | linear (>0) |
| Baseline BMI | GrimAge2 | 3.48 | 9.9 | <1.0E-16 | 11.0 | 10.2% | nonlinear (<0) |
| Baseline BMI | DunedinPAC<br>E | 2.12 | 13.5 | 2.8E-07 | 9.4 | 8.8% | linear (>0) |
| Change in BMI | PAC | 3.06 | 2.1 | 9.7E-02 | 3.6 | 3.6% | None |
| Change in BMI | HPS | 2.67 | 11.9 | 2.5E-07 | 10.3 | 10.3% | nonlinear (<0) |
| Change in BMI | ProtAge | 1.00 | 0.3 | 5.6E-01 | 0.2 | 0.2% | None |
| Change in BMI | Adipose | 2.92 | 10.0 | 9.8E-07 | 9.2 | 9.2% | nonlinear (<0) |
| Change in BMI | Horvath | 1.00 | 0.0 | 9.6E-01 | 0.0 | 0.0% | None |
| Change in BMI | Hannum | 1.00 | 0.0 | 9.9E-01 | 0.2 | 0.2% | None |
| Change in BMI | PhenoAge | 2.59 | 2.7 | 4.4E-02 | 3.5 | 3.5% | nonlinear (<0) |
| Change in BMI | GrimAge2 | 2.94 | 3.4 | 1.0E-02 | 4.5 | 4.5% | None |
| Change in BMI | DunedinPAC<br>E | 1.00 | 24.1 | 1.5E-06 | 6.2 | 6.2% | linear (>0) |
| BMI (wave 5) | PAC | 1.94 | 8.1 | 1.5E-04 | 5.5 | 5.0% | linear (>0) |
| BMI (wave 5) | HPS | 1.05 | 64.6 | <1.0E-16 | 15.8 | 15.5% | linear (>0) |
| BMI (wave 5) | ProtAge | 3.40 | 2.6 | 3.0E-02 | 3.5 | 2.7% | nonlinear (<0) |
| BMI (wave 5) | Adipose | 1.00 | 31.0 | <1.0E-16 | 7.3 | 7.1% | linear (>0) |
| BMI (wave 5) | Horvath | 1.00 | 3.7 | 5.5E-02 | 1.0 | 0.7% | None |
| BMI (wave 5) | Hannum | 1.00 | 3.4 | 6.8E-02 | 0.9 | 0.6% | None |
| BMI (wave 5) | PhenoAge | 1.27 | 7.9 | 1.3E-03 | 3.9 | 3.5% | linear (>0) |
| BMI (wave 5) | GrimAge2 | 1.00 | 25.4 | 1.2E-06 | 7.2 | 6.7% | linear (>0) |
| BMI (wave 5) | DunedinPAC<br>E | 1.00 | 52.7 | <1.0E-16 | 12.4 | 12.1% | linear (>0) |

In quantifying the associations between aging clocks and BMI at wave 5, all associations were significant except for the Hannum and Horvath clocks (Table 2; Figure 1). In contrast to the analyses using baseline BMI, the results did not indicate nonlinearity in the associations with HPS and GrimAge2. However, ProtAge (adjusted for chronological age) was nonlinearly associated with BMI at wave 5, with the graphical representation suggesting that such nonlinearity is likely caused by individuals with BMI values above 30 kg.m^-2^, for whom the ProtAge estimates are higher than average (Figure 1).

**Figure 1:**
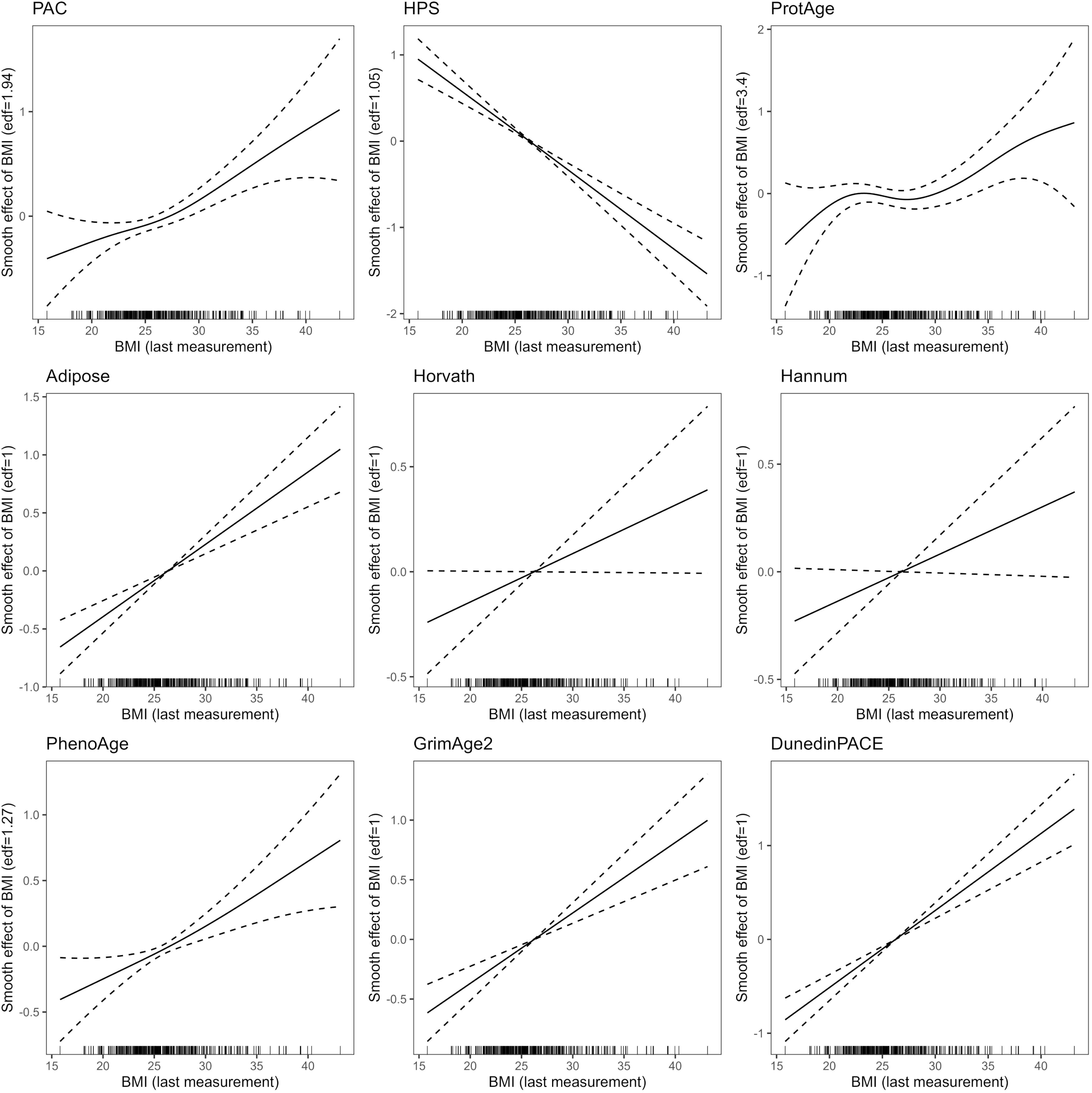
Graphical representation of associations between BMI at wave 5 and biological aging from generalized additive models. Biological age estimates were adjusted for chronological age and scaled. The y-axis represents the effect of BMI on biological aging, which may vary across the range of BMI values (x-axis). The significance of the associations and whether the models suggest that the associations are nonlinear are shown in Table 2.

Biological aging was associated with very long-term changes in BMI for the HPS, Adipose, DunedinPACE, and PhenoAge clocks independent of chronological age and baseline BMI (Table 2; Figure 2). Of these associations, only the DunedinPACE estimate showed no significant improvement in model fit over a simpler linear model, suggesting that such an association can be assumed to be linear. The HPS, Adipose and PhenoAge clocks showed nonlinearity in associations with changes in BMI, with HPS (R^2^=10.3%) and Adipose (R^2^=9.2) clocks being relatively strongly associated with changes in BMI. Nonlinearity in associations was reflected as U-shaped or S-shaped patterns (Figure 2), where associations between changes in BMI and biological aging are likely to be null for individuals who gained close to average units of BMI/year, but significant for individuals who had large increases in their BMI over time. Despite graphical evidence that PAC and GrimAge2 may also be nonlinearly related to changes in BMI, these associations did not reach statistical significance (p=0.09 and p=0.10, respectively).

**Figure 2:**
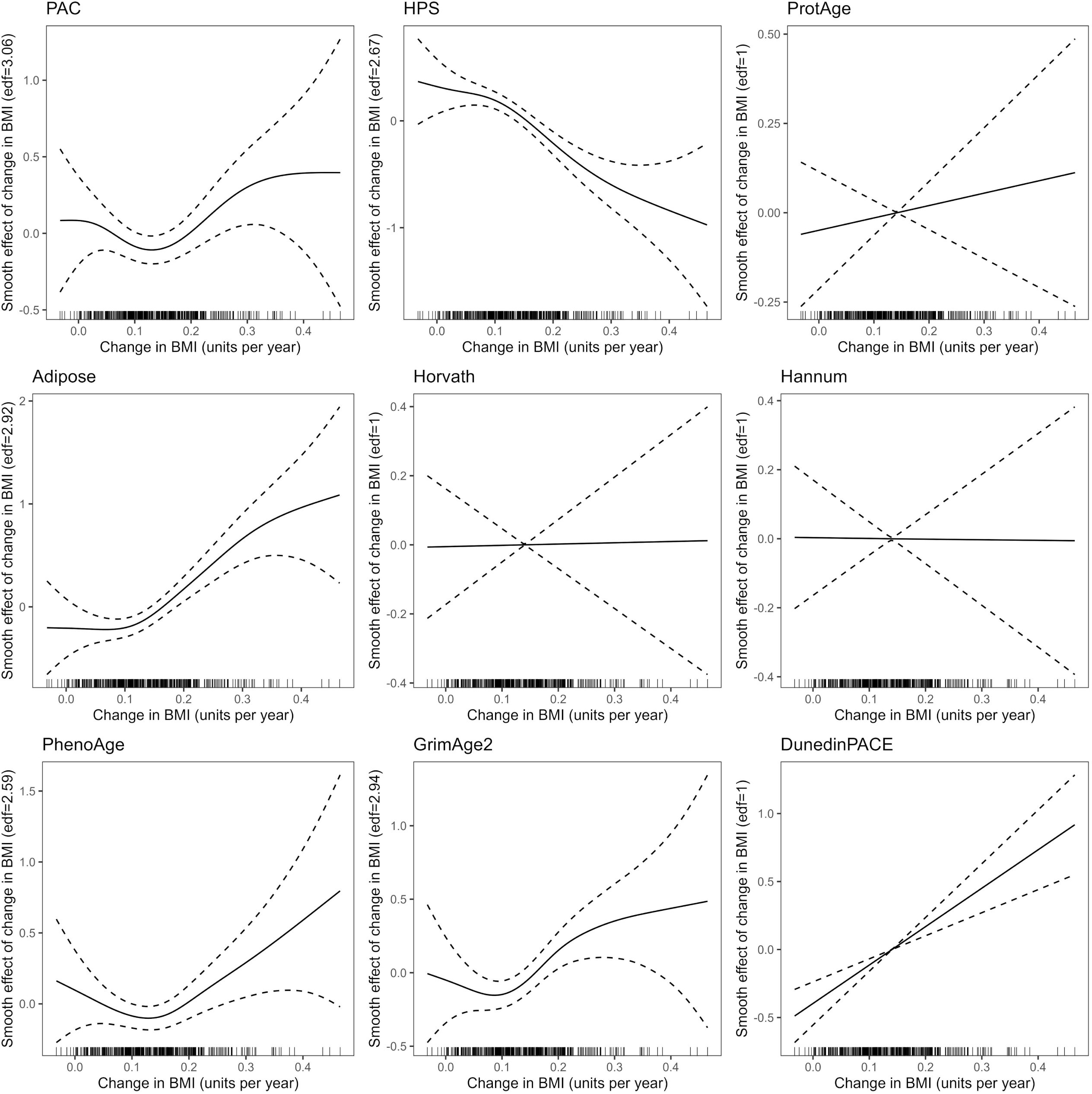
Graphical representation of associations between changes in BMI and biological aging from generalized additive models. Biological age estimates were adjusted for chronological age and scaled. Baseline BMI was added as a covariate in models. The y-axis represents the effect of the change in BMI on biological aging, which may vary across the range of rate of change in BMI values (x-axis). The significance of the associations and whether the models suggest that the associations are nonlinear are shown in Table 2.

### Smooth interactions of baseline BMI with changes in BMI and BMI at blood sampling in explaining biological aging

We examined whether baseline BMI interacted with changes in BMI or BMI at wave 5 in capturing variability in biological aging adjusted for chronological age (Table 3; Figure 3). Joint smooth interactions between baseline BMI and changes in BMI captured most of the variability in the HPS estimate (R^2^=20.7%), which outperformed other clocks by a wide margin (R^2^<14% for all other clocks). The same was true for the joint smooth interaction between baseline BMI and wave 5 BMI, which best captured HPS variability (R^2^=22.7%). This suggests that smooth interactions between baseline BMI with wave 5 BMI and change in BMI are most strongly reflected in the HPS clock. PAC, HPS, Adipose, and PhenoAge showed nonlinearity in the joint interaction between baseline BMI and changes in BMI on biological aging. PAC, HPS, ProtAge, and GrimAge2 showed nonlinearity in the joint interaction between baseline BMI and wave 5 BMI on biological aging.

**Figure 3:**
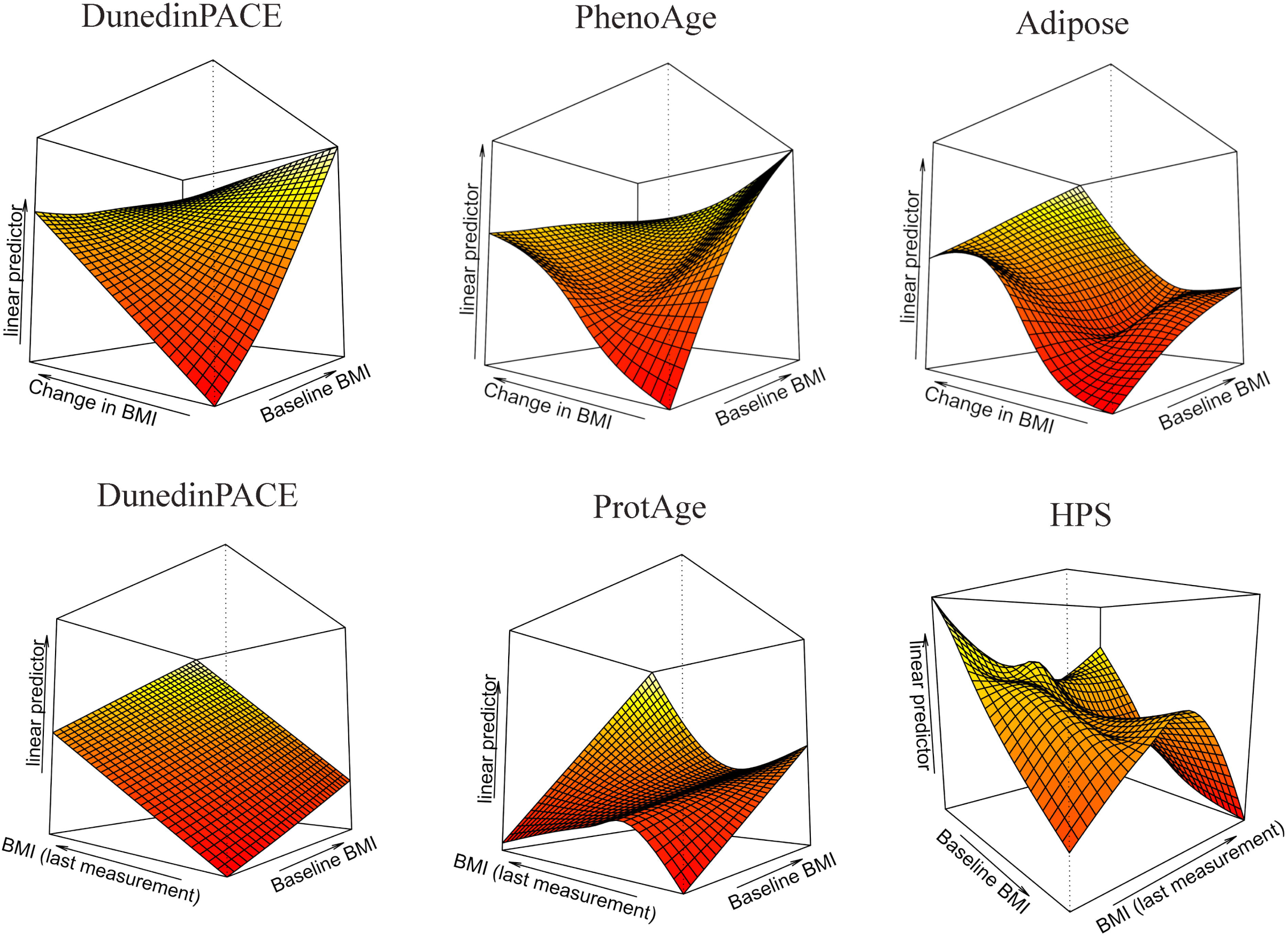
3D graphical representations of joint smooth interactions between baseline BMI with change in BMI and wave 5 BMI in explaining variability in aging clocks. Flat surfaces indicate linearity in the associations between joint smooth interactions and biological aging (adjusted for chronological age), whereas twisted surfaces suggest nonlinearity. DunedinPACE was strongly linearly associated with smooth interaction terms between baseline BMI with change in BMI (top left) and BMI at blood sampling (bottom left) (Table 3). For interactions between baseline BMI and change in BMI, the PhenoAge (top center) and Adipose (top right) clocks showed increasing degrees of nonlinear complexity. For interactions between baseline BMI and wave 5 BMI, ProtAge (bottom center) and HPS (bottom right) showed increasing degrees of nonlinear complexity.

**Table 3:**
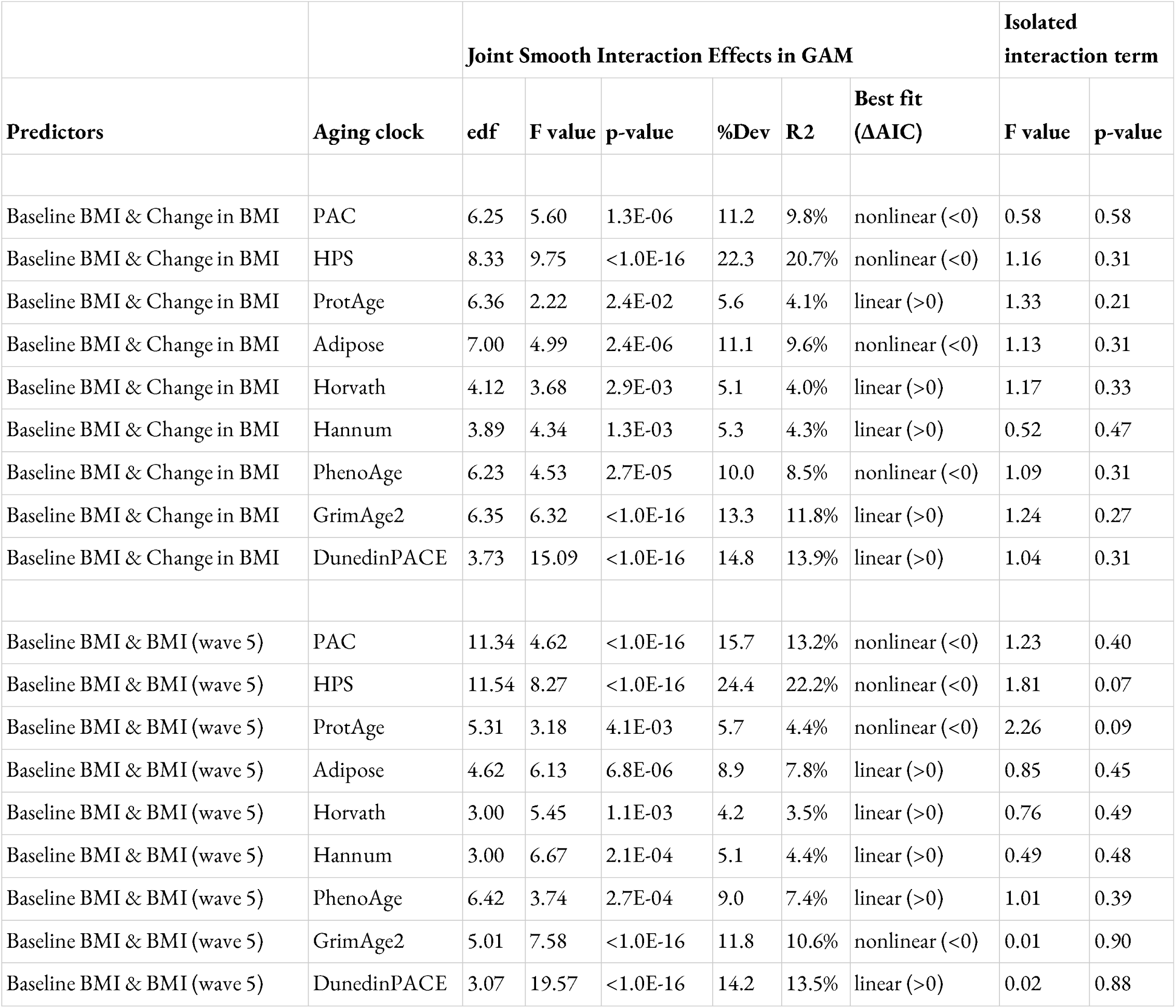
Smooth interactions of baseline BMI with changes in BMI and BMI at blood sampling in explaining biological aging. Aging clock estimates were adjusted for chronological age, and residuals were scaled. GAMs were fitted using either *te()*or *ti()* functions, with the former used to assess the overall smooth interaction in explaining biological aging (and therefore includes the interaction effect as well as the individual effect of both predictors). The *ti()* function was then used to separate the interaction terms and assess the significance of the interaction term independent of the individual effects of the predictors. ΔAIC is defined as the AIC of the nonlinear model minus the AIC of the linear counterpart to indicate whether associations were nonlinear. %Dev: percentage of variation explained. R2: coefficient of determination. edf: effective degrees of freedom.

When we isolated the interaction terms from the individual terms of the predictors in GAMs, no significant results were identified (p-values>0.05) (Table 3). However, for the HPS and ProtAge clocks, we observed suggestive evidence for an interaction between baseline BMI and wave 5 BMI in explaining biological aging (p=0.07 and p=0.09, respectively) (Table 3; Figure 3). This suggests that BMI in young adulthood may modulate the strength of the association between BMI and biological aging at ∼60 years of age, independent of the individual effect of BMI in young adulthood on biological aging itself. However, caution should be exercised in interpreting these results as statistical significance was not reached.

## Discussion

Our study drew on a well-phenotyped sample of twins followed for almost four decades to provide an insightful portrait of how long-term BMI trajectories relate to biological aging. As such, we demonstrate that past BMI assessed 40 years prior to the estimation of biological aging is associated with biological aging, as are changes in BMI over 40 years. Approximately one-third of the significant associations between biological aging and baseline BMI, changes in BMI, and wave 5 BMI (being the BMI at blood sampling) exhibited nonlinear patterns, as indicated by GAMs. This highlights the need to recognize nonlinearity in associations between biological aging and longitudinal body weight. Finally, we observed some evidence for interactions between baseline BMI and wave 5 BMI in capturing biological aging, independent of the individual effects of both predictors. Studies that further examine these interactions in larger samples are encouraged, as additional evidence of interactions would emphasize the need for early prevention of obesity and highlight the long-term burden of high body weight on lifelong aging.

Of the four significant associations identified between biological aging and changes in BMI (adjusted for chronological age and baseline BMI), one was with PhenoAge, which was also shown to be associated with weight gain in Cao et al. [26]. Our study therefore replicates this finding over a longer follow-up and suggests that not only is PhenoAge associated with changes in BMI, but that this association is nonlinear, which was also recently suggested by another study [20]. Li et al. [20] also examined associations between weight change and DunedinPACE where models suggested nonlinearity, whereas in our study such association was shown to be linear. However, the observed nonlinearity of DunedinPACE with weight change reported in Li et al. likely arises because the authors found an association only with weight gain —not with weight loss— with the former following a linear trend, as in our study. The two other clocks we identified associated with long-term changes in BMI were HPS and Adipose, the former being based on a proteomic signature of healthspan. This finding is novel as, to our knowledge, we are the first to examine changes in weight or BMI using proteomic aging clocks. This highlights the need for more studies with different age groups and study designs to include proteomic aging clocks.

While several studies have examined cross-sectional (and to a lesser extent longitudinal) associations of weight or BMI with epigenetic clocks, our study adds a major novelty to the literature since we also investigated proteomic clocks. One assumption that may prevail in the scientific community is that because DNA methylation is strongly imprinted by past lifestyle (such as smoking or weight) [43,44], derived estimates of biological aging may better reflect long-term changes in BMI or the BMI that participants had almost 40 years ago compared to proteomic estimates of biological aging. However, this is not what we observed in the current study; proteomic aging clocks were relatively strongly associated with BMI 40 years ago. Nonlinearity and interactions with baseline BMI observed in associations with proteomic clocks are additional evidence of their ability to reflect past BMI. However, while HPS showed the strongest associations both cross-sectionally and longitudinally with BMI, associations with epigenetic clocks such as GrimAge2 or DunedinPACE were also relatively strong. In addition, large differences were observed between proteomic clocks in their association with BMI trajectories, and the same was observed between the epigenetic clocks.

Consequently, we did not observe an overall superiority of proteomic aging estimates over epigenetic aging estimates in reflecting trajectories of BMI, or *vice vers*.*a*Thus, from an epidemiological perspective, we cannot conclude on the advantages or disadvantages of aging clocks for obesity research based on whether they are based on proteomic or epigenetic data. Instead, our results suggest that the way aging clocks are statistically constructed and trained may be more important than the omics on which they are based in their associations with BMI trajectories. While associations between BMI or changes in BMI and the 1^st^ generation clocks (i.e., clocks trained to predict chronological age) were relatively modest in strength, those with 2^nd^ and 3^rd^ generation clocks (e.g., clocks trained on mortality or healthspan and aging pace) were stronger, regardless of whether the clocks were based on epigenetic or proteomic data. Further studies are needed to confirm or refute this observation, and studies incorporating aging clocks from other omics, such as metabolomics [45], may be of interest to disentangle whether the omics on which aging clocks are built have a substantial influence on how they reflect changes in BMI. Differences between proteomic clocks and epigenetic clocks may lie primarily in their underlying biology, as even though model performance may be comparable at the population level, the clocks do not characterize the same aspect of aging whether defined from proteomic or epigenetic data, as evidenced by weak to moderate pairwise correlations between epigenetic and proteomic clocks after adjustment for chronological age. Thus, shifting from an epidemiological to a biological perspective may exacerbate the differences between proteomic and epigenetic clocks.

Although our study has clear strengths, there are limitations. The first is the sample size, which is relatively modest and may have limited statistical power. This issue may have been especially pronounced when examining interactions between baseline BMI with wave 5 BMI and changes in BMI, where several p-values were found to be relatively low, however not falling below the p=0.05 threshold. However, cohorts with BMI measured over decades in participants with both proteomic and epigenetic data are rare. Another limitation might be the number or the choice of aging clocks used; while the richness of our study lies in the inclusion of nine aging clocks, the development of biological aging clocks has skyrocketed, with novel clocks being now frequently published especially on proteomic data. Thus, our inclusion of proteomic aging clocks is only partial with respect to all the proteomic aging clocks that have been developed in recent months during the preparation of this paper and that may appear later. We focused on four proteomic clocks published (or preprinted) in 2024 and based on UK Biobank data, but studying more clocks, also built from other omics and populations, may provide new insights into how and why changes in BMI translate to increased biological aging. Another limitation is that the aging clocks in the current study differ in their correlations with chronological age, with correlations often lower than those reported in the original publications. We partly attribute these moderate correlations to the relatively homogeneous age range of the EH-Epi sample and the greater variability in biological aging compared to chronological age. Finally, our study focused on BMI, which does not fully reflect obesity status. While our findings are likely to be broadly applicable to obesity research, the biological and social mechanisms underlying obesity, particularly morbid obesity, may differ slightly from those of BMI. The advantage of using BMI is that it is available for large numbers of individuals, but additional studies focusing on obesity status or more accurate markers of body fat may be of even greater public health interest.

In conclusion, our study provides a valuable platform for the scientific community to examine how proteomic aging clocks compare to epigenetic clocks in reflecting long-term changes in BMI and past and current BMI. Assessment of nonlinearity in associations highlights the need to avoid linear modeling in some contexts, and interaction analyses suggest that past BMI, as assessed four decades prior to blood sampling, may modulate associations between biological aging and body weight, which requires additional studies to confirm. Thus, our study highlights the complexity of the relationship between longitudinal body weight and aging and targets early prevention of obesity as a preventive strategy to improve healthy aging.

## Authors’ contributions

GD, MO, and JK conceptualized the study. GD developed the study methodology, performed statistical analyses, prepared figures and tables, and drafted the manuscript. AH processed the methylation data and GD processed the proteomic and anthropometric data. Clock calculations in the EH-Epi sample were performed as follows: AH calculated the epigenetic clocks, AA calculated the ProtAge clock, and GD calculated the PAC, HPS and Adipose clocks. MO and JK supported the creation of the EH-Epi sample and related omics with grants. GD, AA, AH, MO, and JK participated in the discussion and interpretation of the results. All authors were involved in revising the manuscript and approved its final version.

## Funding

Data collection in the Finnish Twin Cohort, including the EH-Epi sample has been supported by the Academy of Finland (Grants 265240, 263278, 308248, 312073, 336832 to JK and 297908 to MO) and the Sigrid Juselius Foundation (to MO).

## Data availability

The Finnish Twin Cohort data used in the analysis is deposited in the Biobank of the Finnish Institute for Health and Welfare (https://thl.fi/en/web/thl-biobank/forresearchers). It is available to researchers after written application and following the relevant Finnish legislation.

## Acknowledgements

We gratefully acknowledge Prof Mark Daly for facilitating the acquisition of the ProtAge clock in the EH-Epi sample.

## Ethics approval and consent to participate

The study protocol was approved by the Institutional Ethics Board of the Hospital District of Helsinki and Uusimaa, Finland (ID 154/13/03/00/11) and the Institutional Review Board of Augusta University.

## Conflicts of Interest

The authors have no conflicts of interest to declare.

## Notes

### Competing Interest Statement

The authors have declared no competing interest.

### Summary of Updates

Affiliations have been fixed; Ethics approval section has been added.

