## supplementary material for "Associations between 40-year trajectories of BMI and proteomic and epigenetic aging clocks: deciphering nonlinearity and interactions"

**Table S1:** Description of the aging clocks used in the current study. **Caption:** All individuals with epigenetic ages also had proteomic ages. N: number of twins with available data.

| Clock | Omic type | N | Original article | Description |
|---|---|---|---|---|
| PAC | Proteomic | 401 | Kuo et al., 2024a | This clock measures biological aging using Gompertz models with >100 blood proteins as predictors. Model training was conducted in a sample of the UK Biobank. |
| HPS | Proteomic | 401 | Kuo et al., 2024b | This clock estimates the 10-year risk of reaching the end of healthspan using Gompertz models with 86 blood proteins and age as predictors. Model training was performed on a sample from the UK Biobank. Unlike other clocks, HPS is negatively correlated with chronological age, as lower values indicate poorer prognosis. |
| ProtAge | Proteomic | 401 | Argentieri et al., 2024 | The ProtAge Clock was designed to predict chronological age from blood proteins, 204 of which were used for its development in the UK Biobank. ProtAge showed strong predictive potential for major chronic diseases. |
| Adipose | Proteomic | 401 | Goeminne et al., 2025 | The Adipose Clock is one of several organ-specific clocks built using blood proteins and elastic net regression in the UK Biobank to predict chronological age (1st generation). This clock is constructed to reflect the biological aging of adipose tissues. |
| Horvath | Epigenetic | 379 | Horvath, 2013 | This pan-tissue clock estimates biological age based on DNA methylation patterns that best estimate chronological age across the genome. It is widely used to estimate biological age in various tissues, providing a general measure of aging. |
| Hannum | Epigenetic | 379 | Hannum et al., 2013 | This clock uses DNA methylation to estimate chronological age in blood samples. It primarily estimates chronological age and correlates with mortality risk and age-related health outcomes. |
| PhenoAge | Epigenetic | 379 | Levine et al., 2018 | This clock is based on a composite of clinical biomarkers of phenotypic age, which is predicted from DNA methylation patterns. It estimates mortality risk and captures an individual's physiological health and functional capacity. |
| GrimAge2 | Epigenetic | 379 | Lu et al., 2022 | This clock is built on nine DNAm-based surrogates of plasma proteins, DNAm PACKYRS, age and sex to estimate mortality risk. It predicts lifespan, mortality risk and the likelihood of developing age-related diseases. |
| DunedinPACE | Epigenetic | 379 | Belsky et al., 2022 | This clock was trained on longitudinal physiological measurements of the pace of aging captured by DNA methylation at specific CpG sites. This clock predicts how fast an individual is aging relative to their chronological age, providing insight into future risk of age-related decline. |

### Characteristics of the biological clocks

The mean square error (MSE) for the ProtAge, Adipose, Horvath, Hannum, PhenoAge, and GrimAge2 clocks was 30.94, 41.18, 20.10, 54.59, 68.56, and 28.50, respectively. The Mean Absolute Error (MAE) for the ProtAge, Adipose, Horvath, Hannum, PhenoAge, and GrimAge2 watches was 4.90, 5.33, 3.45, 5.99, 6.91, and 4.19, respectively. Epigenetic aging estimates, originally calculated in a larger sample of twins with a wide age range, correlated highly with chronological age in the Finnish Twin Cohort (ChronoAge) (Pearson r>0.90), except for DunedinPACE, which is a measure of the pace of aging and thus does not correlate with ChronoAge. Correlations with ChronoAge in the EH-Epi sample, for which the age range is lower, are presented below in addition to those of the proteomic clocks. Pairwise correlations between biological age clocks corrected for chronological age and scatter plots of biological age estimates versus chronological age are shown below as well.

**Table S2:** Pairwise correlations between nine biological aging clocks and chronological age (designated here as ChronoAge).

| | PAC | HPS | ProtAge | Adipose | Horvath | Hannum | PhenoAge | GrimAge2 | DunedinPACE | ChronoAge |
|---|---|---|---|---|---|---|---|---|---|---|
| PAC | 1 | -0.84 | 0.58 | 0.21 | 0.36 | 0.38 | 0.48 | 0.57 | 0.4 | 0.48 |
| HPS | -0.84 | 1 | -0.45 | -0.3 | -0.37 | -0.36 | -0.47 | -0.64 | -0.51 | -0.38 |
| ProtAge | 0.58 | -0.45 | 1 | 0.15 | 0.44 | 0.43 | 0.45 | 0.31 | 0.11 | 0.77 |
| Adipose | 0.21 | -0.3 | 0.15 | 1 | 0.08 | 0.07 | 0.11 | 0.1 | 0.09 | 0.11 |
| Horvath | 0.36 | -0.37 | 0.44 | 0.08 | 1 | 0.84 | 0.67 | 0.41 | 0.26 | 0.56 |
| Hannum | 0.38 | -0.36 | 0.43 | 0.07 | 0.84 | 1 | 0.76 | 0.53 | 0.37 | 0.52 |
| PhenoAge | 0.48 | -0.47 | 0.45 | 0.11 | 0.67 | 0.76 | 1 | 0.59 | 0.43 | 0.5 |
| GrimAge2 | 0.57 | -0.64 | 0.31 | 0.1 | 0.41 | 0.53 | 0.59 | 1 | 0.7 | 0.37 |
| DunedinPACE | 0.4 | -0.51 | 0.11 | 0.09 | 0.26 | 0.37 | 0.43 | 0.7 | 1 | 0 |
| ChronoAge | 0.48 | -0.38 | 0.77 | 0.11 | 0.56 | 0.52 | 0.5 | 0.37 | 0 | 1 |

**Table S3:** Pairwise correlations between biological aging clocks adjusted for chronological age.

| | PAC | HPS | ProtAge | Adipose | Horvath | Hannum | PhenoAge | GrimAge2 | DunedinPACE |
|---|---|---|---|---|---|---|---|---|---|
| PAC | 1 | -0.81 | 0.38 | 0.18 | 0.12 | 0.18 | 0.28 | 0.49 | 0.46 |
| HPS | -0.81 | 1 | -0.27 | -0.28 | -0.2 | -0.21 | -0.29 | -0.58 | -0.56 |
| ProtAge | 0.38 | -0.27 | 1 | 0.1 | 0.03 | 0.07 | 0.11 | 0.04 | 0.17 |
| Adipose | 0.18 | -0.28 | 0.1 | 1 | 0.03 | 0.01 | 0.07 | 0.07 | 0.09 |

| | | | | | | | | | |
|---|---|---|---|---|---|---|---|---|---|
| Horvath | 0.12 | -0.2 | 0.03 | 0.03 | 1 | 0.78 | 0.49 | 0.26 | 0.31 |
| Hannum | 0.18 | -0.21 | 0.07 | 0.01 | 0.78 | 1 | 0.8 | 0.43 | 0.42 |
| PhenoAge | 0.28 | -0.29 | 0.11 | 0.07 | 0.49 | 0.8 | 1 | 0.54 | 0.56 |
| GrimAge2 | 0.49 | -0.58 | 0.04 | 0.07 | 0.26 | 0.43 | 0.54 | 1 | 0.75 |
| DunedinPACE | 0.46 | -0.56 | 0.17 | 0.09 | 0.31 | 0.42 | 0.56 | 0.75 | 1 |

**Figure S1:** Individual body mass index trajectories over the follow-up period (on the left side) with the number of measurements per person given on the right-side panel

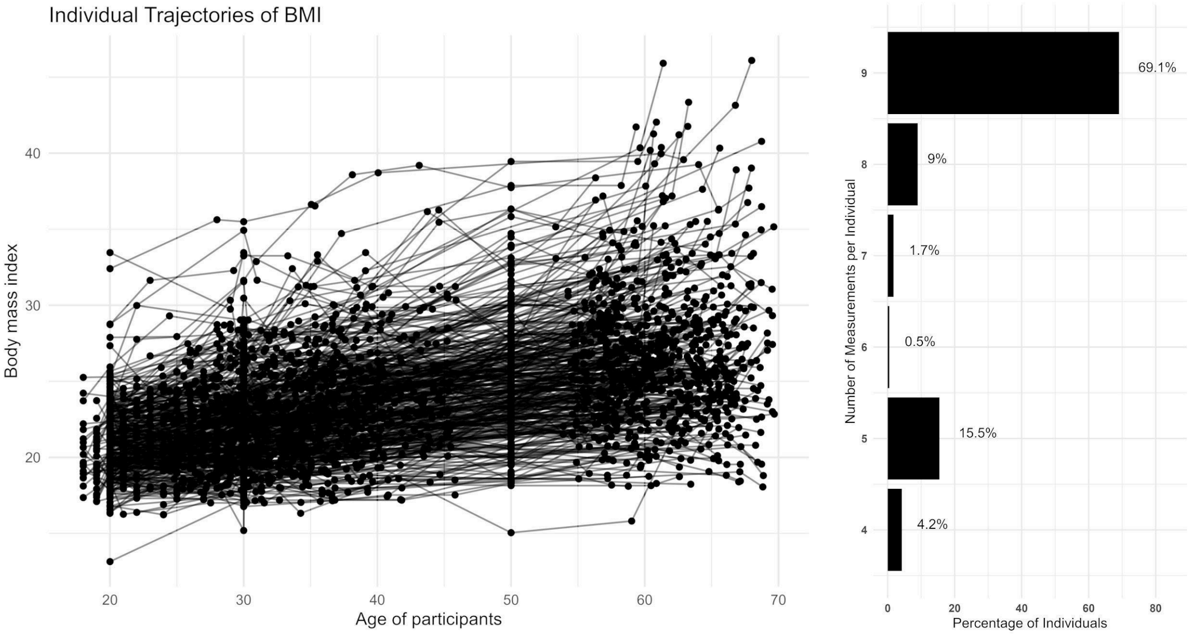

**Figure S2:** Scatter plots of biological age vs chronological age. HPS and DunedinPACE are not shown because their values do not characterize estimates of biological age, but rather related measures (healthspan for HPS and pace of aging for DunedinPACE).

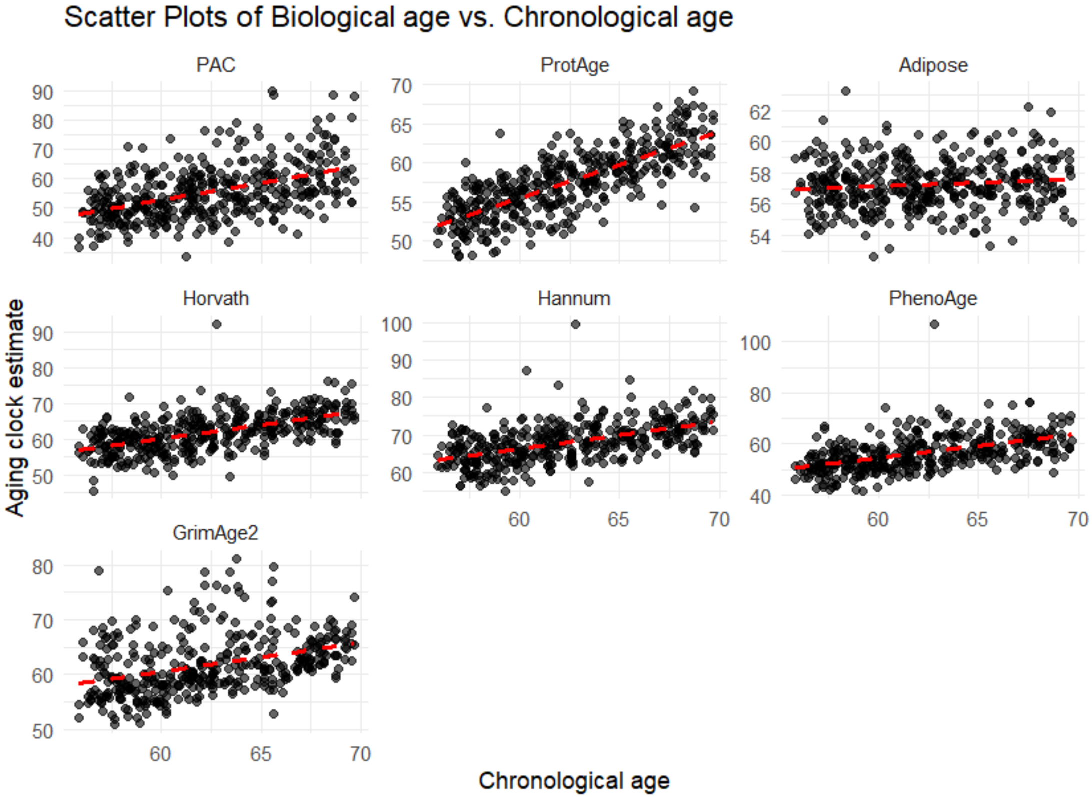

**Figure S3:** Graphical representation of associations between baseline BMI and biological aging from generalized additive models. **Caption:** Aging clock estimates were adjusted for chronological age and scaled. The y-axis represents the effect of baseline BMI on biological aging, which may vary across the range of baseline BMI values (x-axis). The significance of the associations and whether the models suggest that the associations are nonlinear are shown in Table 2.

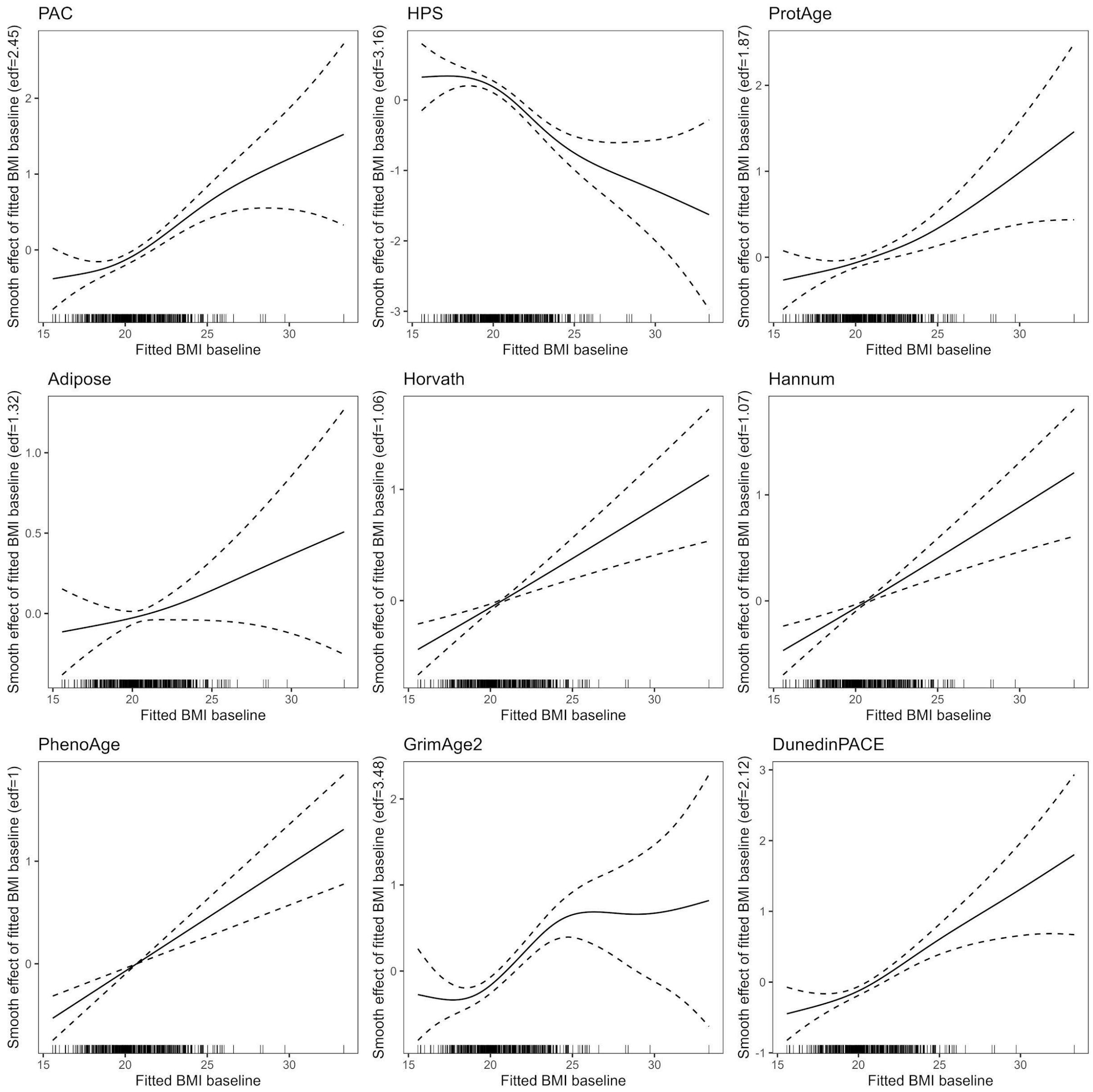
